# Automated Segmentation of Trunk Musculature with a Deep CNN Trained from Sparse Annotations in Radiation Therapy Patients with Metastatic Spine Disease

**DOI:** 10.1101/2025.01.13.25319967

**Authors:** Vy Hong, Steve Pieper, Joanna James, Dennis E Anderson, Csaba Pinter, Yi Shuen Chang, Bulent Aslan, David Kozono, Patrick F Doyle, Sarah Caplan, Heejoo Kang, Tracy Balboni, Alexander Spektor, Mai Anh Huynh, Mario Keko, Ron Kikinis, David B Hackney, Ron N Alkalay

## Abstract

**Purpose:** Given the high prevalence of vertebral fractures post-radiotherapy in patients with metastatic spine disease, accurate and rapid muscle segmentation could support efforts to quantify muscular changes due to disease or treatment and enable biomechanical modeling for assessments of vertebral loading to improve personalized evaluation of vertebral fracture risk. This study presents a deep-learning approach for segmenting the complete volume of the trunk muscles from clinical CT images trained using sparsely annotated data.

**Materials and Methods:** we extracted 2,009 axial CT images at the midpoint of each vertebral level (T4 to L4) from clinical CT of 148 cancer patients. The key extensor and flexor muscles (up to 8 muscles per side) were manually contoured and labeled per image in the thoracic and lumbar regions. We first trained a 2D nnU-Net deep-learning model on these labels to segment key extensor and flexor muscles. Using these sparse annotations per spine, we trained the model to segment each muscle’s entire 3D volume

**Results:** The proposed method achieved comparable performance to manual segmentations, as assessed by expert radiologists, with a mean Dice score above 0.769. Significantly, the model drastically reduced segmentation time, from 4.3-6.5 hours for manual segmentation of 14 single axial CT images to approximately 1 minute for segmenting the complete thoracic-abdominal 3D volume.

**Conclusion:** The approach demonstrates high potential for automating 3D muscle segmentation, significantly reducing the manual intervention required for generating musculoskeletal models, and could be instrumental in enhancing clinical decision-making and patient care in radiation oncology.

**Summary:** A deep learning 2D nnU-Net model, trained on a sparse set of 2D muscle annotations, successfully segmented the entire volume of 20 thoracolumbar muscles from cancer patients’ clinical CT data. The model showed a remarkable increase in segmentation efficacy and generalizability, achieving comparable performance to manual segmentations in delineating each muscle anatomy.

**Key Points:**

▪ A deep learning model (2D nnU-Net), developed using a sparse set of single axial CT-slice at each mid-per vertebral level, containing manual image annotation of 20 thoracic and lumbar muscles, achieved comparable performance to manual segmentations, as assessed by expert radiologists, with a mean Dice score above 0.769.
▪ The model drastically reduced segmentation time, from 4.3-6.5 hours for manual segmentation of 14 single axial CT images to approximately 1 minute for segmenting the complete thoracic-abdominal 3D volume.
▪ Radiologist assessment based on a Likert scale (0-5) for clinical acceptability of the muscle anatomical segmentation showed strong model performance for a representative sample of clinical CT data (a (mean(SD) of 4.66 (0.73)) and external data (4.66 (0.73).

## 1. Introduction

In patients treated with palliative radiation therapy for metastatic spine disease, up to 40% suffer clinically significant vertebral fracture (VF) [1]. Evaluating VF risk is critical for managing these patients [2]. Biomechanically, VF may occur if the vertebral loading, largely produced by the thoracic and abdominal muscles during daily activities [3], exceeds the lesioned vertebrae strength, causing a Load-Strength-Ratio (LSR) value higher than one [4]. A recent musculoskeletal simulation study using subject-specific muscle morphology and spinal curvature found osteolytic vertebrae with significantly higher LSR and osteosclerotic vertebrae with significantly lower LSR than values computed for a cohort of 250 cancer-free age- and sex-matched subjects from the Framingham Heart Study [5]. These results suggest that bone metastasis-mediated changes in LSR affect spinal stability and VF risk, a finding in agreement with clinical reports [6]. The lack of efficient, automated 3D segmentation of abdominal and thoracic musculature from clinical imaging has rendered prohibitive the application of these simulations to estimate the roles of LSR and patients’ daily activities in defining VF risk in clinical studies.

Patient-specific musculoskeletal simulations allow insight into the relative magnitudes of vertebral loading conditions (compressive, moment-based) that cannot be measured non-invasively in patients [7]. Such models can be improved by using detailed geometry and other properties of the thoracic and lumbar musculature and osseous spine calculated at each vertebral level [7]. No comprehensive individual muscle segmentation data is available for cancer patients, and manual segmentation is impractical for creating personalized simulations for each patient. Deep learning (DL) methods employing 2D U-Net models [8–11] achieved 0.95 agreement with expert segmentation for L3 abdominal musculature. 3D segmentation is harder due to data scarcity. Applying 2D U-Net successively [12] had 82.8% agreement for erector spinae. However, these approaches depend on scarce high-quality manual segmentations. Self-supervised and semi-supervised methods leverage limited data [13–15] but target single lumbar slices unsuitable for musculoskeletal modeling.

Our goal was to adapt, apply, and validate a DL algorithm for efficient, automated segmentation of thoracoabdominal musculature from sparse clinical CT data, focusing on extensor and flexor muscles used in spinal musculoskeletal mechanics [16]. To achieve this goal, we trained a DL model using detailed single-slice segmentations, had expert radiologists evaluate the anatomical accuracy of each muscle segmentation, compared DL and manual segmentation for axial CT data, evaluated the model’s performance on external data outside the study training/validation dataset, and tested its generalizability using public CT data.

## 2. Methods

### 2.1 Cancer Study Participants

This study comprised 148 patients (49 females and 99 males, Table 1) receiving radiotherapy (RT) between September 2020 and July 2023. The patients recruited under NIH grant were previously consented to the research project. Inclusion criteria included 1) receiving RT for solid tumor metastasis in the bony thoracic or lumbar spine. 2) had no prior RT to the same vertebral level, no surgery with hardware to the same or adjacent vertebral level(s), no kyphoplasty or vertebroplasty to the same vertebral level(s), and did not have diseases of abnormal bone metabolism such as Paget’s or Cushing’s disease.

**Table 1.**
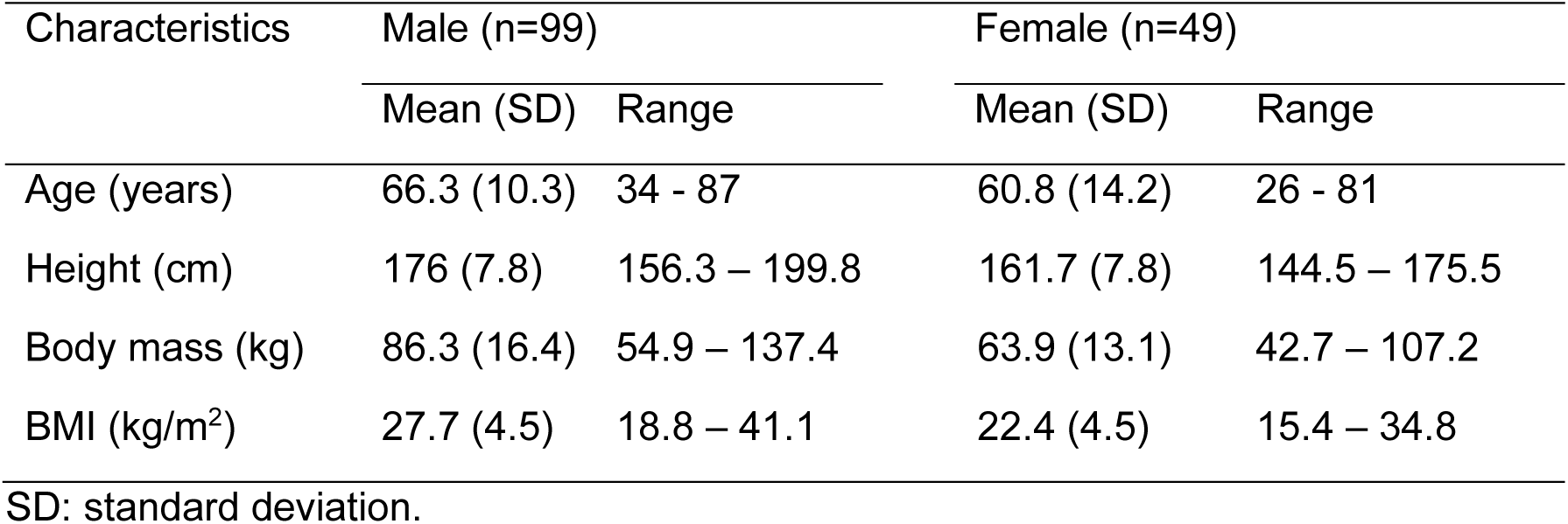
Demographic characteristics of the cancer patients recruited as part of NIH grant R01AR075964.

| Characteristics | Male (n=99) |  | Female (n=49) |  |
| --- | --- | --- | --- | --- |
|  | Mean (SD) | Range | Mean (SD) | Range |
| Age (years) | 66.3 (10.3) | 34 - 87 | 60.8 (14.2) | 26 - 81 |
| Height (cm) | 176 (7.8) | 156.3 – 199.8 | 161.7 (7.8) | 144.5 – 175.5 |
| Body mass (kg) | 86.3 (16.4) | 54.9 – 137.4 | 63.9 (13.1) | 42.7 – 107.2 |
| BMI (kg/m<sup>2</sup>) | 27.7 (4.5) | 18.8 – 41.1 | 22.4 (4.5) | 15.4 – 34.8 |

SD: standard deviation.

### 2.2 Computer Tomography Data

We obtained the CT data as part of the standard protocol for simulation for our study patient’s radiotherapy treatment planning. Simulation details and CT Image acquisition protocols are detailed in supplement S.1. Images were de-identified using the anonymization feature in MIM (version 7.1.12, MIM Software Inc, Cleveland, OH) and further anonymized by study staff who removed all dates from scans.

### 2.3. Ground-truth muscle annotations

Following published protocol [16], ground truth manual muscle segmentation was performed in Analyze^TM^ (Biomedical Imaging Resource, Mayo Clinic, Rochester, MN) [17] by a single research associate specially trained for this task. A detailed description of the process and training is provided in Supplement S.2. In total, we extracted 2,009 axial CT image data slices from the 148 CT volumes corresponding to the midpoint of each vertebral level, and individual muscles were contoured and labeled per level (Table 2). Figure 1 illustrates a manual segmentation and associated labels performed for L3 and T5 levels, with Table 2 detailing the muscle segmented per vertebral level. Each manual muscle segmentation required 2-4 minutes per muscle (initial tracing and required corrections), requiring 20-30 min segmentation per axial CT slice (both left and right, 14 muscles on average), leading to segmentation times of 4.3-6.5 hours per subject for 13 axial CT slices.

**Table 2.** lists the thoracic and lumbar muscles measured from axial CT scans at each vertebral level.

| Muscle | T4 | T5 | T6 | T7 | T8 | T9 | T10 | T11 | T12 | L1 | L2 | L3 | L4 | L5 |
| --- | --- | --- | --- | --- | --- | --- | --- | --- | --- | --- | --- | --- | --- | --- |
| Pectoralis Major | X | X | X | X | X | X |  |  |  |  |  |  |  |  |
| Rectus Abdominis |  |  |  |  |  |  | X | X | X | X | X | X | X | X |
| Serratus Anterior | X | X | X | X | X | X | X | X |  |  |  |  |  |  |
| Trapezius | X | X | X | X | X | X | X | X |  |  |  |  |  |  |
| Latissimus Dorsi | X | X | X | X | X | X | X | X | X | X | X | X |  |  |
| External Oblique |  |  |  |  |  |  | X | X | X | X | X | X | X | X |
| Internal Oblique |  |  |  |  |  |  |  |  |  |  | X | X | X | X |
| Psoas Major |  |  |  |  |  |  |  |  |  | X | X | X | X | X |
| Erector Spinae | X | X | X | X | X | X | X | X | X | X | X | X | X | X |
| Transversospinalis | X | X | X | X | X | X | X | X | X | X | X | X | X | X |
| Quadratus<br>Lumborum |  |  |  |  |  |  |  |  |  | X | X | X | X |  |

**Figure [1].**
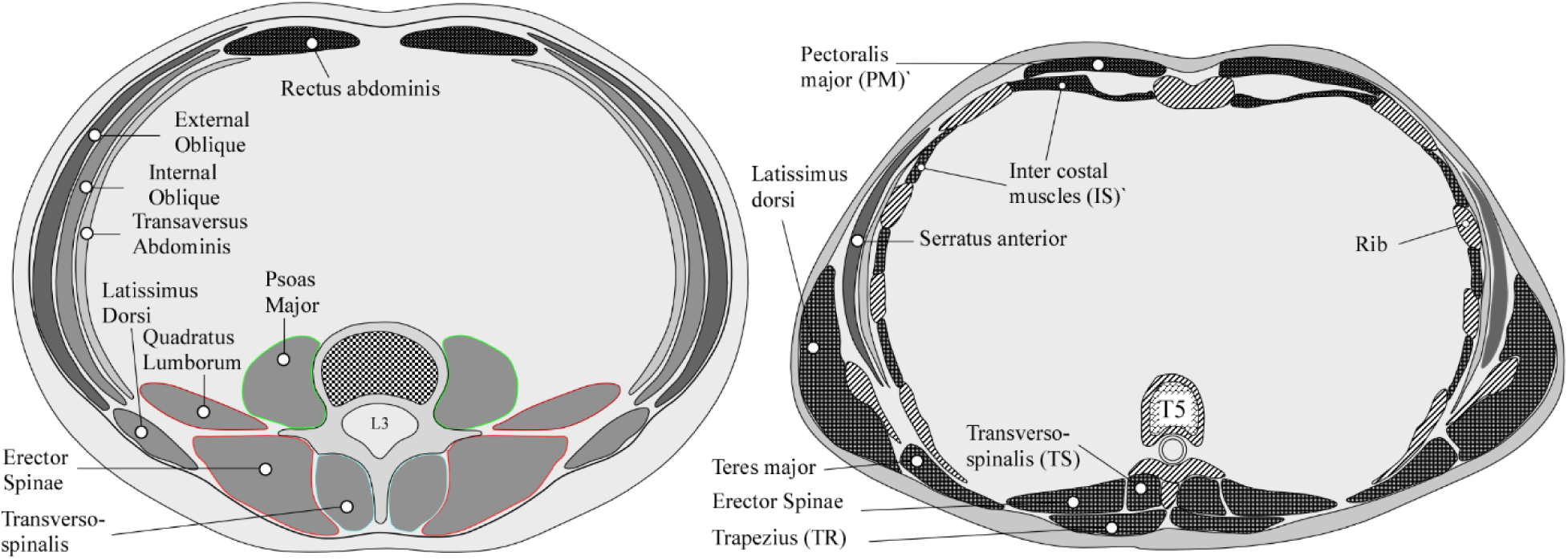
Graphic illustration (RNA) of the muscle groups segmented in the lumbar and thoracic regions.

### 2.4 Establishing deep learning muscle segmentation

To build an efficient, fully automated pipeline for accurate segmentation of the thoracic, thoracolumbar and lumbar, we employed a two-stage approach:

1. 2D segmentation: we utilized the nnU-Net v2 package from the German Cancer Research Center (DKFZ) without modifications for training and inference (Isensee, Jaeger et al. 2021). We trained the nnU-Net exclusively on mid-vertebral 2D slices corresponding to manually-segmented slices as ground truth. We treat these slices as representative samples of the whole upper body, allowing the model to learn meaningful features without requiring densely annotated 3D volumes; we hypothesized that the resulting model would generalize and be able to segment CT slices that are not at the mid-vertebral levels. Data preparation followed the nnU-Net preprocessing pipeline, and model planning and training were conducted using the 2D nnU-Net configuration. Figure (2) illustrates the initial iteration of the model. We adopted the default 5-fold training method, where one-fifth of the training data was held out for validation in each fold. Model training and validation were performed on the Jetstream2 cloud computing environment (https://jetstream-cloud.org/) at Indiana University [18, 19].
2. 2D to 3D: The 3D thoracic and abdominal muscle segmentation was performed by successively applying the trained 2D model to the stack of axial CT image data within the CT volume with the complete 3D segmentation (Figure 2), requiring on average 1 minute in to process all slices. The default data augmentation for training the nnU-Net model includes flipping the images left-right and front-back. However, we found that this was causing the model to determine the left/right orientation of the segmented muscles incorrectly. Upon further review, we retrained the model without any left-right or front-back flipping during data augmentation. This eliminated the issue of the model incorrectly labeling the laterality of the muscles in the segmentation outputs.

**Figure [2].**
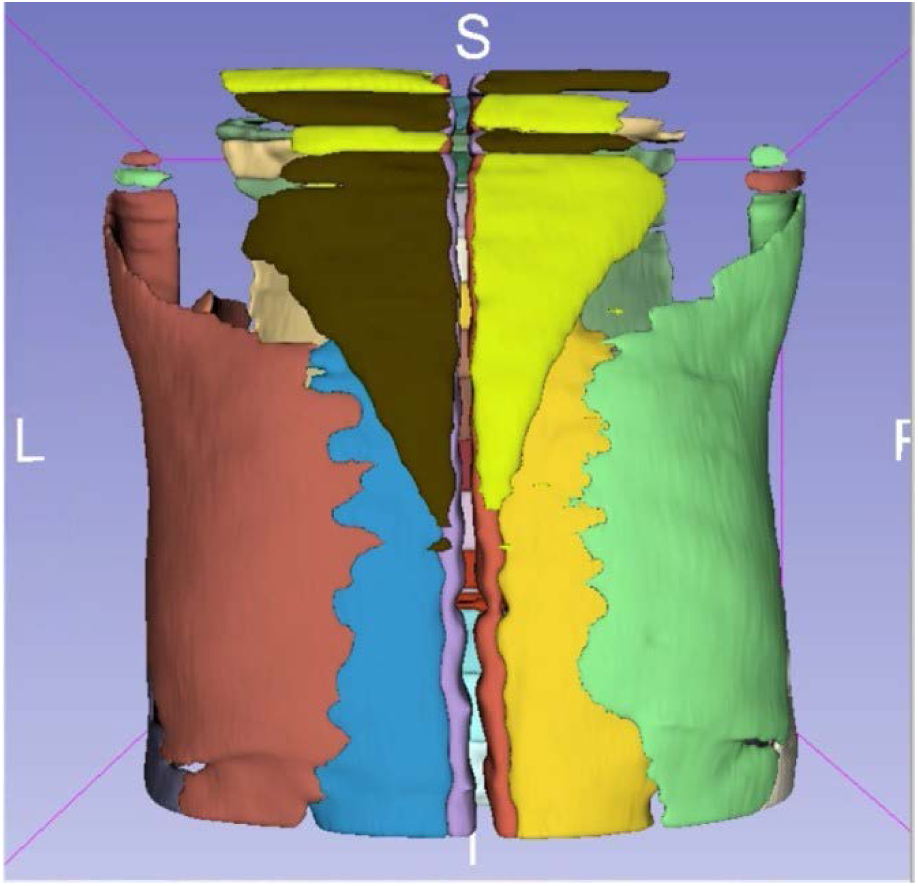
Initial result where the algorithm yielded multiple muscles laterality flipped near the top of the volume, such as Trapezius (Yellow / Black).

### 2.5 Model performance

Two experienced radiologists evaluated the quality of manual- and model-generated muscle segmentations (Table 2), blind to whether the segmentations were generated by manual or deep-learning. The evaluation followed a Likert scale (0-5) for clinical acceptability applied to axial CT data from the following groups: (1, n=30) Manual- and (2, n=30) DL-segmentation, selected from the training data, (3, n=30) randomly selected from the full 3D muscle segmentation and (4) DL segmentation applied to external data selected from the National Lung Screening Trial (NLST) [20], supplement S.3.

### 2.6 Data analysis

Radiologists’ ratings were averaged for each CT image and muscle left and right segmentation to create a single average rating score in each CT dataset (Manual-, DL-segmentation, Randomly selected, and External data). In turn, the left and right sides were collapsed to a single array of scores. We computed each muscle’s grand mean and corresponding standard deviation per evaluation and evaluation group (Manual-, DL-segmentation, Randomly selected CT and External data). Mann-Whitney test was used to compare the difference in rating between Manual and DL muscle segmentation (Group 1 vs. 2) with a significance level of 0.05.

## Code Availability

All codes and processing scripts are freely available at https://github.com/Spine-Biomechanics-Group-Alkalay-Lab/Spine-Muscle-Segmenter.git.

## 3. Results

### 3.1 DL-muscle segmentation

Using a single g3.xl Jetstream2 instance (A100, 40GB video RAM, https://jetstream-cloud.org/), model training took seven days, with the nnU-Net achieving a mean Dice score across five folds greater than 0.769. Training took approximately one minute per epoch and ran for 2,000 epochs per fold, stabilizing quickly with subsequent epochs adding little to the result, Figure [3.A]. Once training was completed, the nnU-Net model inference applied on a slice-by-slice approach required, on average, 1 minute per CT volume to generate a continuous 3D segmentation of the upper body muscles, Figure 4.

**Figure 3A]:**
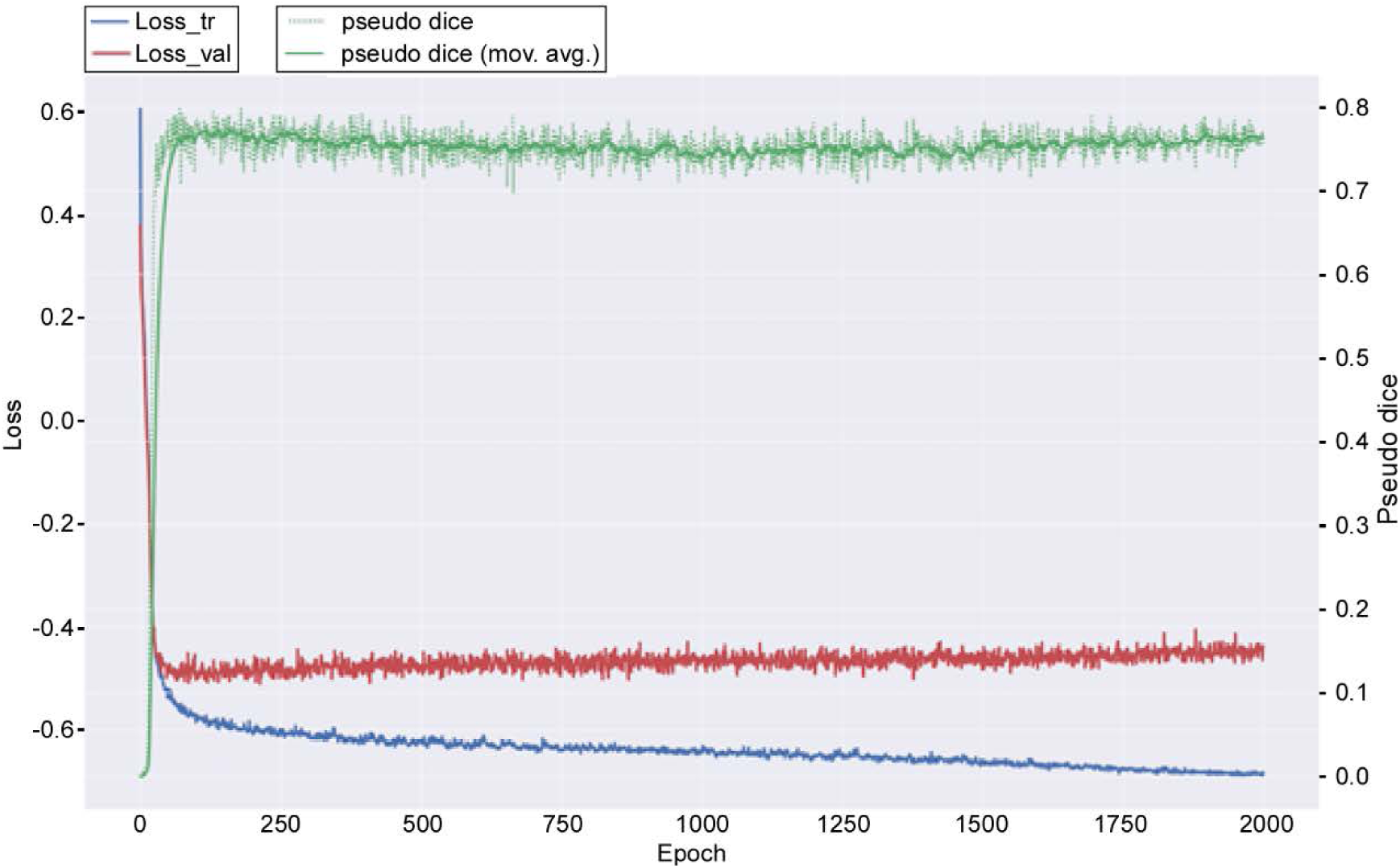
The training process for fold 0 is typical of the results across all 5 folds. A graphic comparison of the manual and DL segmentation for an axial slice is presented in Fig 3.B

**Figure 3B].**
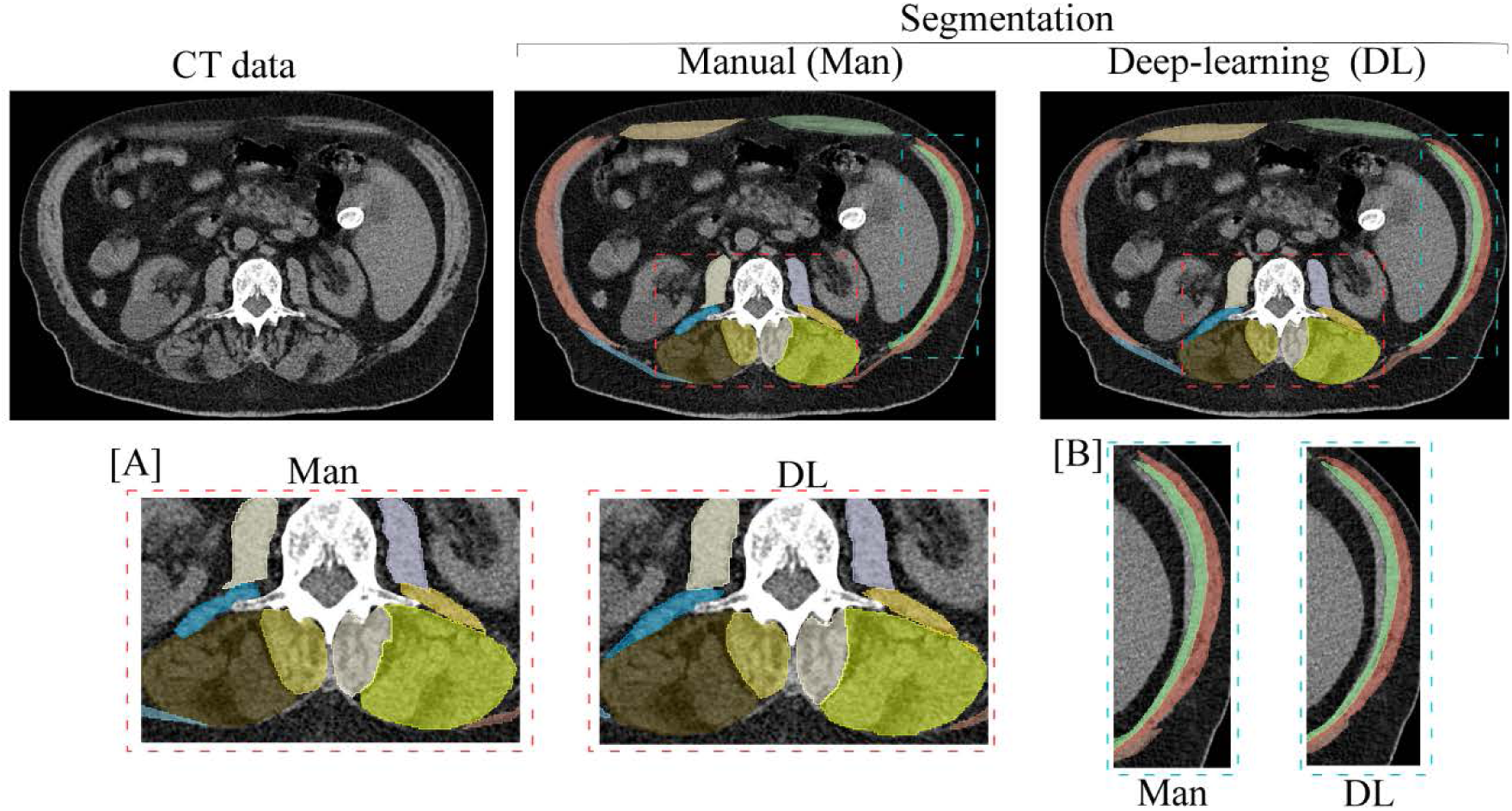
Manual and deep learning segmentation for axial CT data. The magnified sections compare the DL segmentation quality for the extensor [A] and oblique [B] musculature.

**Figure [4].**
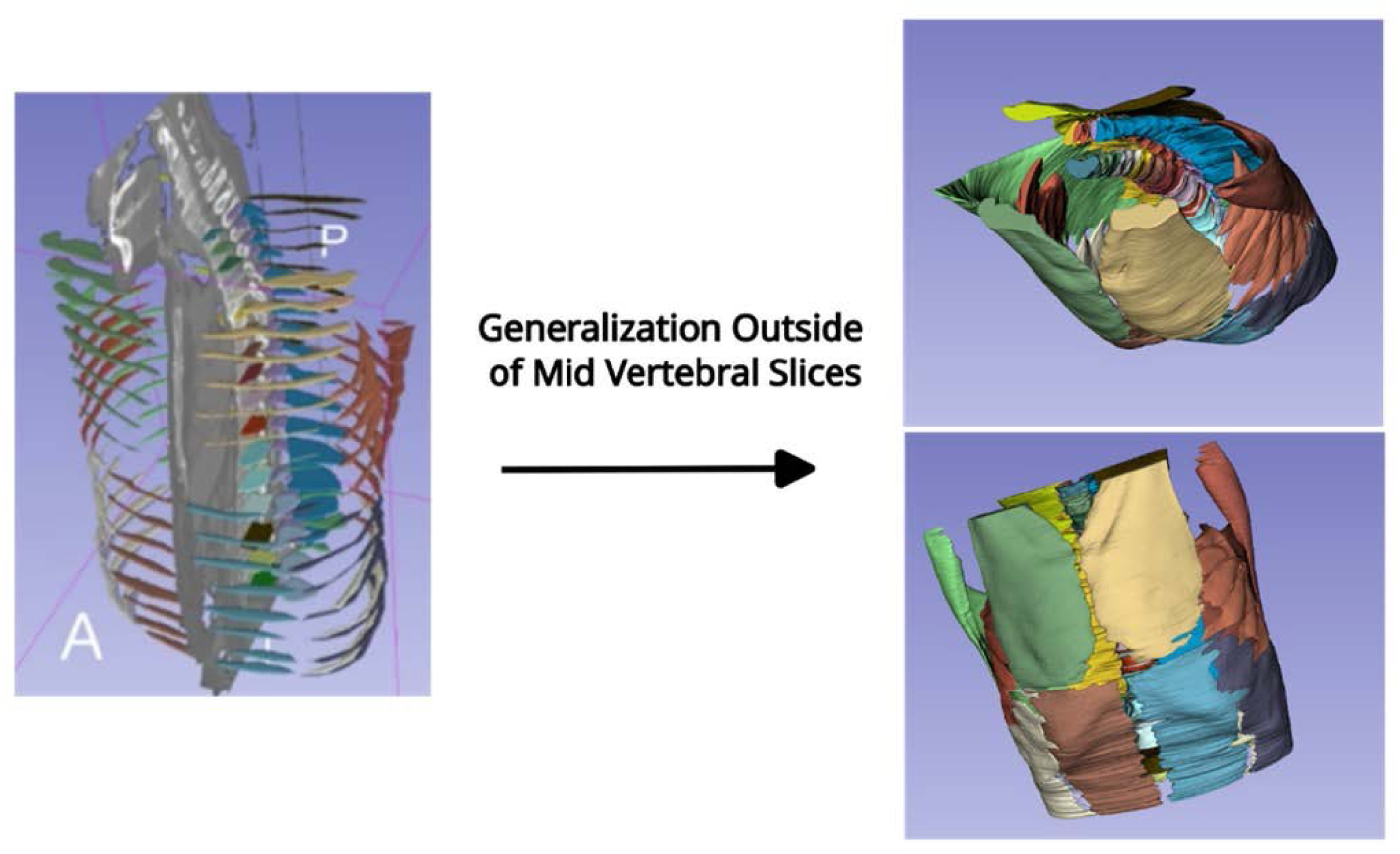
shows the resulting volume when inferring every slice of the volume. Horizontal banding, visible in the abdominal region as seen in the frontal view figure (lower right), is believed to be due to breathing artifacts during the CT acquisition but may merit further investigation. Please note that as no breath hold control was performed, the banding in the abdominal muscles is likely due to breathing.

### 3.2 Model validation

a. *Manual vs. DL muscle segmentation*: We first evaluated the nnU-Net model segmentation ability to delineate individual muscle anatomy with manual segmentation based on the radiologist scores. Mann-Whitney test analysis found no statistically significant difference in radiologist ratings between the manual (Group 1, supplement S3) and DL-generated (Group 2, supplement S3) muscle segmentation for each muscle comparison, Table 3. Although the individual comparisons were not statistically significant, manual and DL segmentations for the Erector Spinae were lower than the group mean score, with a mean difference of -3.7% and 9.0%, respectively (Table 3). Plotting the radiologist scores by level score (Figure 5) suggested that the reduced scores occurred predominantly at the higher thoracic levels.
b. *3D and out-of-study muscle segmentations*: The radiologist’s review scores for random samples of muscle segmentation extracted from the 3D muscle segmentation volumes (Group 3, supplement S3) and the external data (Group 4, supplement S3) are summarized in Table 4. In group 3, 65.7% of the average segmentation quality scores were equal to or exceeded 4.6. For group 4, 64.3% of the segmentation quality scores were equal to or exceeded 4.6. The lowest review scores correspond to the Psoas Major muscle, with a score of 4.12 in group 4.

**Figure [5].**
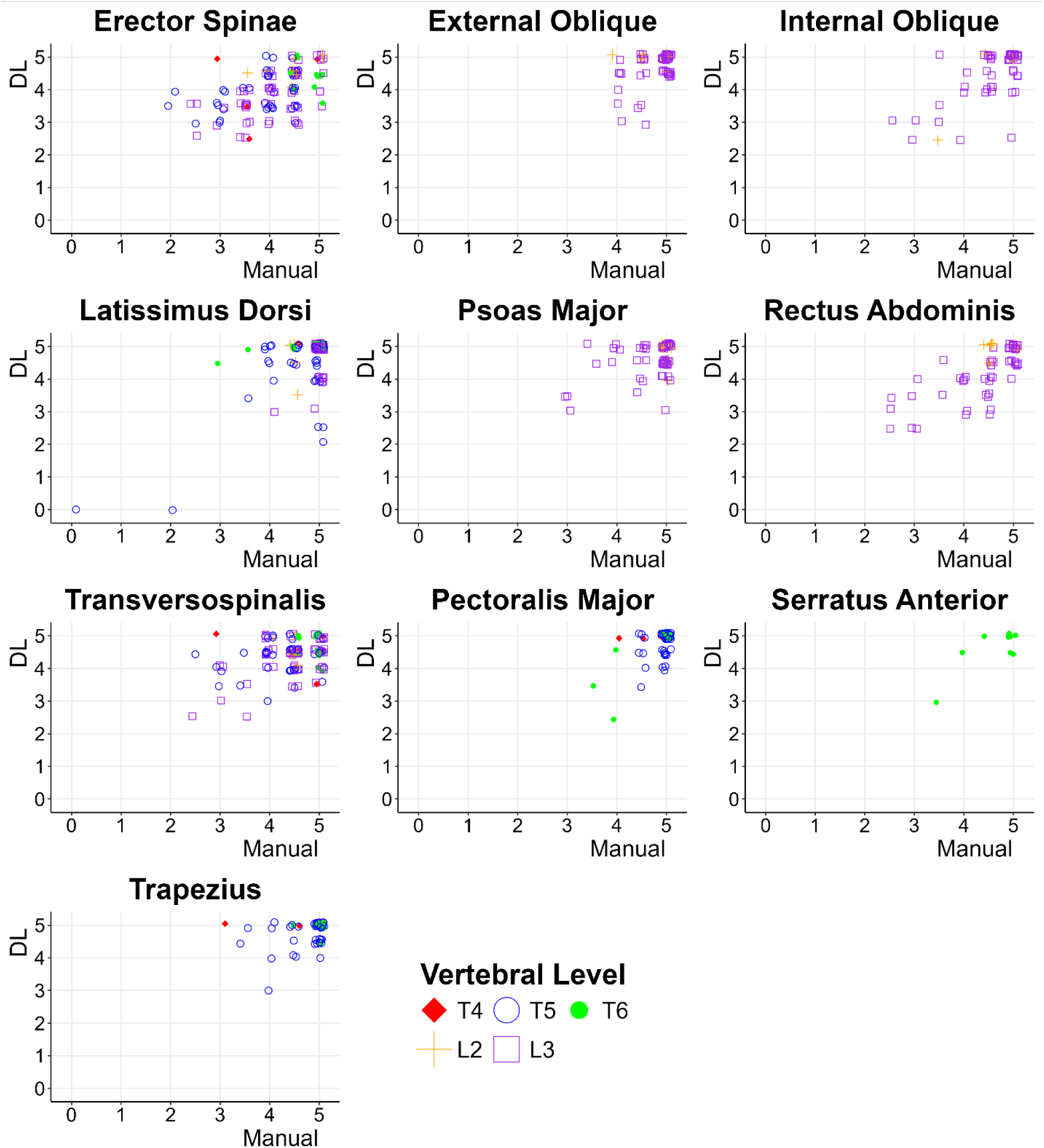
presents scatter plots of radiologist assessment scores of the manual and DL segmentation by muscle and vertebral level. The radiologist scores found that segmentation accuracy was not uniform across spinal muscles and was less accurate at upper thoracic levels. We added minimal jitter to the marker position within each sub-figure to enhance clarity and readability.

**Figure [6].**
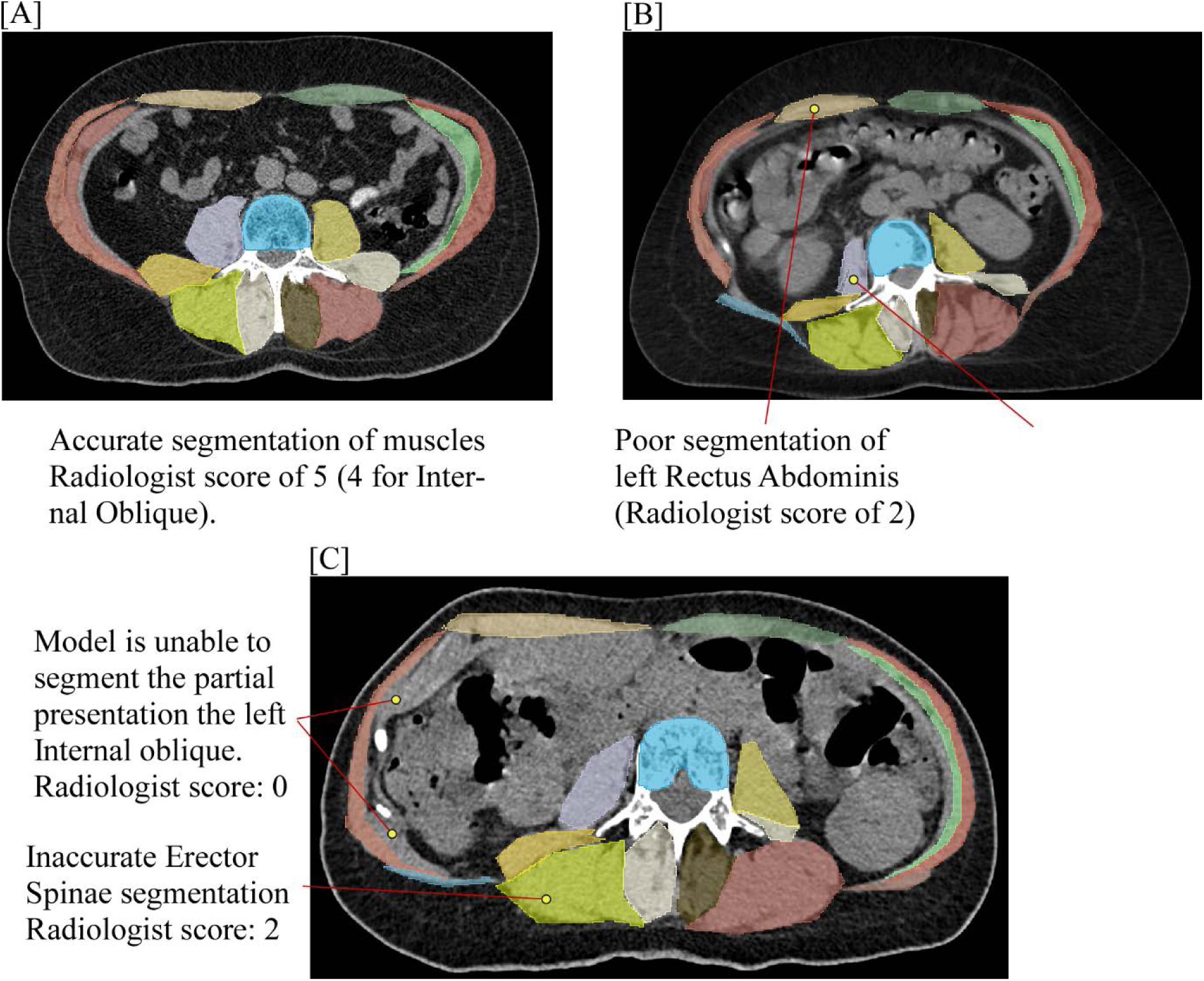
Graphic presentation of a successful DL muscle segmentation [A]), corresponding to a high score by the radiologist raters and low scores by the radiologist rater resulting from either poor [B] or partial and missing segmentation [C].

## 4. Discussion

This study successfully employed and validated a DL model for efficient, automated thoracic and lumbar musculature segmentation from sparsely annotated clinical CT data, focusing on the main extensor and flexor muscles used in spinal musculoskeletal simulations. Based on radiologist assessment scores, the 2D nnU-Net approach showed no statistically significant differences between manual and DL segmentations. However, this performance was not uniform across muscles, with challenges in segmenting thin muscles at higher thoracic levels. The model greatly improved segmentation efficiency compared to a human operator. Testing this level of inference performance in an independent set of subjects suggests the model’s generalizability.

**Table 3.** Summary statistics (mean, standard deviation) for the manual and corresponding DL segmentations of the cancer patient data (groups 1, manual segmentation, and 2, DL segmentation.

| Muscle segmented | Rating |  |  |
| --- | --- | --- | --- |
|  | Group 1: | Group 2: |  |
|  | Manual segmentation* | DL Segmentation* | P value** |
| **Erector Spinae** | 4.09 (0.71), n=120 | 4.00 (0.67), n=120 | 0.219 |
| **External Oblique** | 4.76 (0.36), n=60 | 4.70 (0.50), n=60 | 0.756 |
| **Internal Oblique** | 4.57 (0.61), n=56 | 4.46 (0.79), n=56 | 0.837 |
| **Latissimus Dorsi** | 4.75 (0.65), n=114 | 4.61 (0.86), n=114 | 0.409 |
| **Pectoralis Major** | 4.86 (0.32), n=60 | 4.73 (0.53), n=60 | 0.192 |
| **Psoas Major** | 4.68 (0.55), n=60 | 4.58 (0.52), n=60 | 0.091 |
| **Rectus Abdominis** | 4.44 (0.72), n=61 | 4.28 (0.76), n=61 | 0.216 |
| **Serratus Anterior** | 4.75 (0.50), n=12 | 4.75 (0.50), n=12 | 0.773 |
| **Transversospinalis** | 4.41 (0.59), n=120 | 4.38 (0.52), n=120 | 0.286 |
| **Trapezius** | 4.78 (0.44), n=60 | 4.79 (0.38), n=60 | 0.683 |
| **Grand mean** | 4.55 (0.64), n=716 | 4.46 (0.69), n=716 | 0.012 |

Each muscle's sides (right, left) are collapsed to produce one score.\* The radiologist rating of each segmentation method is presented as mean (standard deviation), n = number of individual muscles rated. \*\* P value from Mann-Whitney test. Description of Groups 1 and 2 are detailed in Supplement S.3.

**Table 4:** Average Radiologist rating for cancer patients, group 3, and external CT database of subjects, group 4.

| **Muscle** | **Group 3** | **Group 4** |
| --- | --- | --- |
|  | **Random Sample \*** | **External Data\*** |
| **Erector Spinae** | 4.60 (0.50), n=120 | 4.65 (0.43), n=120 |
| **External Oblique** | 4.70 (0.70), n=62 | 4.84 (0.34), n=59 |
| **Internal Oblique** | 4.58 (0.84), n=49 | 4.62 (0.25), n=4 |
| **Latissimus Dorsi** | 4.56 (1.17), n=73 | 4.55 (0.81), n=104 |
| **Pectoralis Major** | 4.65 (0.90), n=57 | 4.83 (0.46), n=61 |
| **Psoas Major** | 4.66 (0.92), n=59 | 4.12 (1.24), n=45 |
| **Rectus Abdominis** | 4.71 (0.72), n=62 | 4.78 (0.36), n=60 |
| **Serratus Anterior** | 4.67 (0.56), n=26 | 4.67 (0.56), n=26 |
| **Transversospinalis** | 4.68 (0.46), n=120 | 4.71 (0.45), n=120 |
| **Trapezius** | 4.83 (0.50), n=60 | 4.88 (0.40), n=60 |
| **Grand mean** | 4.66 (0.73), n=688 | 4.66 (0.73), n=688 |

Each muscle's sides (right, left) are collapsed to produce one score.\* The radiologist rating of each segmentation method is presented as mean (standard deviation), n = number of individual muscles rated. Description of Groups 3 and 4 are detailed in Supplement S.3.

Evaluation of muscle composition and mass from clinical CT imaging [21] and the detailed segmentation required for patient-specific musculoskeletal simulations [7] have traditionally relied on manual or semiautomated techniques [22, 23]. These methods are time-consuming [24], prone to error and intra- and interobserver variability [25] and require input from clinical experts with specialized knowledge. Deep learning models achieved an agreement of up to 0.95 with manual-based muscle segmentation at the L3 [8–10]. However, these studies remain directed at assessing cancer cachexia, producing single segmentation of the entire musculature within a single axial CT slice, data unsuitable for creating spinal musculoskeletal models [26]. A nnU-Net model [27] recently demonstrated high-performance automated segmentation of 104 organs [28] within the trunk from clinical CT data. Although providing detailed segmentation of the spinal skeleton, the model provides limited spinal muscle segmentations (Trapezius, Serratus_Anterior and pectoralis_minor). Significantly, this model’s low resolution (1.5mm isotropic) limits its ability to segment the boundaries between thin spinal muscles ((such as external and internal obliques act to stabilize the trunk), affects grouping the spinal extensor muscles (Erector Spinae and Transversospinalis: support forward bending moment imposed by external loading) and the Psoas Major (hip flexor and trunk rotation muscle).

Training on limited annotations per vertebral level to segment the entire 3D volume proved effective, with the model achieving 94% of the average segmentation quality equal to or exceeding the manual segmentation score of 4.6 out of possible 5. Significantly, when applied to external CT data, the model’s strong segmentation performance suggested that it can generalize from training on only a few slices to automated segmentation of the entire musculature. Integrated into a musculoskeletal simulation pipeline, this model’s performance, one minute for automated muscle segmentation of 835 high-resolution axial CT images (0.6mm slice thickness) compared to 4.3-6.5 hours for manual muscle segmentation of 13 axial CT slices (132 hours projected for equivalent patient volume (400, 1.25mm slice thickness CT slices), will allow us, for the first time, to perform high-throughput studies in large cohorts of cancer patients. These patients frequently experience cachexia, characterized by weight loss with the concomitant loss of muscle and/or fat mass, causing functional impairment, high patient frailty and risk of falls [29], and decreased survival [26]. However, despite these well-characterized functional impairments, the role of these muscle changes in affecting cancer patients’ VF risk remains understudied. The ability to comprehensively segment the entire thoracic and lumbar spine muscles will permit large-scale studies of subjects with cancer and other diseases resulting in muscle loss, fostering new insight into their effect on the role of daily spinal loads in affecting the patient VF risk and developing approaches to mitigate this risk.

This study had several limitations. The muscles labeled for this study lacked the transversus abdominis as the segmentation was strictly limited to muscles required for the musculoskeletal model of spinal loading [5]. Although applying the same approach to MR images of the torso would be appealing, we have not assessed performance using this modality. A prior study of MR-based lumbar paraspinal muscle segmentation and subsequent modeling [30] showed that lumbar loading estimates of models created by automatic vs. manual segmentation were correlated. However, that agreement varied by vertebral level, from L5 (R = 0.55) to L2 (R = 0.87). While not yet tested similarly, the current DL muscle segmentation is better-suited to creating musculoskeletal models as it incorporates all key trunk muscles across the full thoracolumbar spine. We acknowledge that there is no true gold standard for these segmentations, ultimately relying on expert subjective evaluation. Ultimately, it is unclear what constitutes a more reliable standard. However, the strong agreement between the automated and manual techniques suggests they should be of comparable accuracy.

In conclusion, this CT DL muscle segmentation model achieved segmentation performance of spinal muscle anatomy comparable to human raters with remarkably higher efficiency, resulting in automated segmentation of the complete volume of human thoracic and lumbar regions. This work represents a significant step toward automated musculoskeletal modeling in cancer patients, potentially enabling routine assessment of vertebral fracture risk in clinical settings and may have applications in other fields of medicine. This advancement could help identify high-risk patients earlier, allowing for more timely interventions and improved patient outcomes.

## Data Availability

All data produced in the present study are available upon reasonable request to the authors.

https://github.com/Spine-Biomechanics-Group-Alkalay-Lab/Spine-Muscle-Segmenter.git

## References

1. Faruqi, S., et al., Vertebral Compression Fracture After Spine Stereotactic Body Radiation Therapy: A Review of the Pathophysiology and Risk Factors. Neurosurgery, 2018. 83(3): p. 314–322.

2. Yao, A., et al., Contemporary spinal oncology treatment paradigms and outcomes for metastatic tumors to the spine: A systematic review of breast, prostate, renal, and lung metastases. J Clin Neurosci, 2017. 41: p. 11–23.

3. Arjmand, N. and A. Shirazi-Adl, Model and in vivo studies on human trunk load partitioning and stability in isometric forward flexions. J Biomech, 2006. 39(3): p. 510–21.

4. Mokhtarzadeh, H. and D.E. Anderson, The Role of Trunk Musculature in Osteoporotic Vertebral Fractures: Implications for Prediction, Prevention, and Management. Curr Osteoporos Rep, 2016. 14(3): p. 67–76.

5. Anderson, D.E., et al., Evaluation of Load-To-Strength Ratios in Metastatic Vertebrae and Comparison With Age- and Sex-Matched Healthy Individuals. Front Bioeng Biotechnol, 2022. 10: p. 866970.

6. Weber, M.H., et al., Instability and impending instability of the thoracolumbar spine in patients with spinal metastases: a systematic review. Int J Oncol, 2011. 38(11): p. 5–12.

7. Bruno, A.G., M.L. Bouxsein, and D.E. Anderson, Development and Validation of a Musculoskeletal Model of the Fully Articulated Thoracolumbar Spine and Rib Cage. J Biomech Eng, 2015. 137(8): p. 081003.

8. Hemke, R., et al., Deep learning for automated segmentation of pelvic muscles, fat, and bone from CT studies for body composition assessment. Skeletal Radiol, 2020. 49(3): p. 387–395.

9. Edwards, K., et al., Abdominal muscle segmentation from CT using a convolutional neural network. Proc SPIE Int Soc Opt Eng, 2020. 11317.

10. Ackermans, L., et al., Deep Learning Automated Segmentation for Muscle and Adipose Tissue from Abdominal Computed Tomography in Polytrauma Patients. Sensors (Basel), 2021. 21(6).

11. Shen, H., et al., A deep learning model based on the attention mechanism for automatic segmentation of abdominal muscle and fat for body composition assessment. Quant Imaging Med Surg, 2023. 13(3): p. 1384–1398.

12. Kamiya, N., Muscle Segmentation for Orthopedic Interventions. Adv Exp Med Biol, 2018. 1093: p. 81–91.

13. Ouassit, Y., et al., A Brief Survey on Weakly Supervised Semantic Segmentation. Int. J. Online Biomed. Eng., 2022. 18: p. 83–113.

14. Cai, H., et al. 3D Medical Image Segmentation with Sparse Annotation via Cross-Teaching between 3D and 2D Networks. in International Conference on Medical Image Computing and Computer-Assisted Intervention. 2023.

15. Çiçek, Ö., et al. 3D U-Net: Learning Dense Volumetric Segmentation from Sparse Annotation. in International Conference on Medical Image Computing and Computer-Assisted Intervention. 2016.

16. Allaire, B.T., et al., Dependence of trunk muscle size and position on age, height, and weight in a multi-ethnic cohort of middle-aged and older men and women. J Biomech, 2023. 157: p. 111710.

17. Robb, R.A., The biomedical imaging resource at Mayo Clinic. IEEE Trans Med Imaging, 2001. 20(9): p. 854–67.

18. Boerner, T., et al., ACCESS: Advancing Innovation: NSF’s Advanced Cyberinfrastructure Coordination Ecosystem: Services & Support. 2023. 173–176.

19. Hancock, D., et al., Jetstream2: Accelerating cloud computing via Jetstream. 2021. 1–8.

20. (ACRIN), L.S.S.g.L.a.t.A.C.o.R.I.N., *National Lung Screening Trial (NLST)* N.C.I. (NCI), Editor. 2009.

21. Fernandez-Fewell, G.D. and M. Meredith, Facilitation of mating behavior in male hamsters by LHRH and AcLHRH5-10: interaction with the vomeronasal system. Physiol Behav, 1995. 57(2): p. 213–21.

22. Olson, B., et al., Establishment and Validation of Pre-Therapy Cervical Vertebrae Muscle Quantification as a Prognostic Marker of Sarcopenia in Patients With Head and Neck Cancer. Front Oncol, 2022. 12: p. 812159.

23. Swartz, J.E., et al., Feasibility of using head and neck CT imaging to assess skeletal muscle mass in head and neck cancer patients. Oral Oncol, 2016. 62: p. 28–33.

24. Ackermans, L., et al., Clinical evaluation of automated segmentation for body composition analysis on abdominal L3 CT slices in polytrauma patients. Injury, 2022. 53 Suppl 3: p. S30–S41.

25. Perthen, J.E., et al., Intra- and interobserver variability in skeletal muscle measurements using computed tomography images. Eur J Radiol, 2018. 109: p. 142–146.

26. Mortellaro, S., et al., Quantitative and Qualitative Radiological Assessment of Sarcopenia and Cachexia in Cancer Patients: A Systematic Review. J Pers Med, 2024. 14(3).

27. Isensee, F., et al., nnU-Net: a self-configuring method for deep learning-based biomedical image segmentation. Nat Methods, 2021. 18(2): p. 203–211.

28. Wasserthal, J., et al., TotalSegmentator: Robust Segmentation of 104 Anatomic Structures in CT Images. Radiol Artif Intell, 2023. 5(5): p. e230024.

29. Morris, R. and A. Lewis, Falls and Cancer. Clin Oncol (R Coll Radiol), 2020. 32(9): p. 569–578.

30. Hess, M., et al., Deep Learning for Multi-Tissue Segmentation and Fully Automatic Personalized Biomechanical Models from BACPAC Clinical Lumbar Spine MRI. Pain Medicine, 2022. 24(Supplement_1): p. S139–S148.

