## Supplementary material for "Automated Segmentation of Trunk Musculature with a Deep CNN Trained from Sparse Annotations in Radiation Therapy Patients with Metastatic Spine Disease": Supplamental data

Suplamental Material

**S.1	CT Image acquisition protocols.**

The CT data was obtained as part of the clinical CT simulation for radiotherapy treatment planning. The Simulations were performed using the Siemens SOMATOM Confidence (Siemens Healthcare GmbH, Erlangen, Germany) or the GE Lightspeed (General Electric Medical System, Waukesha, WI) CT scanner. The CT scans were not gated, and breath-hold techniques were not used. The CT images in the collected datasets had a section thickness of 0.5 or 1.25mm. Table S.1 details the simulation scan parameters.

**Table S.1.**  NIH cancer study Computer Tomography imaging protocol parameters.

| **Radiotherapy** | **CT scanner** | | | |
| --- | --- | --- | --- | --- |
|  | **Siemens SOMATOM Confidence** | | **GE Lightspeed** | |
| Protocol parameters | SBRT | All Others | SBRT | All Others |
| Tube voltage (kVp) | 120 | 120 | 120 | 120 |
| mA | 240-300 | 240-300 | 300 | 300 |
| FOV | A, B | A, B | A, B | A, B |
| Slice Thickness | 0.5mm | 1.25mm | 1.25mm | 1.25mm |
| In-Plane Pixel Size | 0.70-0.98mm | 0.70-0.98mm | 0.70-0.98mm | 0.70-0.98mm |
| Gantry rotation | 1s | 1s | 1s | 1s |
| Gating | None | None | None | None |
| Breath Hold | None | None | None | None |

A: 16cm FOV and B: Skin-to-Skin FOV

**S.2**	**Ground-truth muscle annotations**.

Following published protocol [1], ground truth manual muscle segmentation was performed in Analyze^TM^ (Biomedical Imaging Resource, Mayo Clinic, Rochester, MN) [2] by a single research associate, specially trained for this task via observations, guided analysis, and review of unsupervised segmentation of 12 training scan volumes, which were visually reviewed by an experienced user and compared to existing gold-standard segmentations [1]. These manual segmentations have excellent inter- and intra-rater reliability for both muscle cross-sectional area (CSA) and location relative to the vertebral body, with intra-class correlation coefficients ranging from 0.84 to 0.99 Axial CT image slices corresponding to the centroid of T4 to L4 vertebral bodies were identified and processed using a Sigma filter [3] to reduce noise while preserving the edges of the muscle's tissue boundaries. The Object Extractor module was used to manually trace individual muscle and vertebral body boundaries at each vertebral level. The major spinal flexor and extensor muscles were segmented depending on the CT image ﬁeld of view (FOV) and level viewed (Table 2). Figure 1 illustrates a manual segmentation and associated labels performed for L3 and T5 levels, with Table 2 detailing the muscle segmented per vertebral level.

For this study, 2,009 axial CT image data slices were extracted from the 148 CT volumes corresponding to the midpoint of each vertebral level, and individual muscles were contoured and labeled per level (Table 2). Each manual muscle segmentation required 2-4 minutes per muscle (initial tracing and required corrections), requiring 20-30 min segmentation per axial CT slice (both left and right, 14 muscles on average), leading to segmentation times of 4.3-6.5 hours per subject for 13 axial CT slices. Using a hydroxyapatite phantom scanned asynchronously on each CT system before the patient's simulation CT (Image Analysis, Inc., Lexington, KY, USA), the CT image was standardized, and each muscle contour was processed to exclude voxels outside the 50 to 150 HU range removing voxels associated with pure fat, tendon, and bone from each muscle contour's margin [Johannesdottir et al. 2018]. This asynchronous scan was required due to strict limitations on adding a phantom to the cancer patient's radiotherapy planning CT. The resulting data file containing binary labels for each vertebral level (each muscle cross-sectional area (CSA) and the corresponding vertebral body CSA) was exported in Analyze^TM^ 7.5 format.

**S.3**	**Model performance assessment**.

Two experienced radiologists individually evaluated the quality of model-generated segmentation for each muscle contour (Table 2), blind to whether the CT slice was generated by manual or deep-learning segmentation. This evaluation followed a 0 to 5 Likert scale for clinical acceptability for muscle segmentation contour with 0: Segmentation is unacceptable; 1: Poor (<50% matches muscle anatomy); 2: Inadequate: <75% matches muscle anatomy). 3: Acceptable: >75% but <90% matches muscle anatomy), 4: Good: small differences from the muscle anatomy) with 5: Best: Completely matches the muscle anatomy. This scale was applied to the following groups:

1) *Group 1: Manual-segmentation:* We selected 30 CT images from thoracic [T4: n=2), T5: n=22 and T6; n=6] and lumbar (L2: n=4, L3: n=26] levels, yielding 757 individual muscle segmentations.

2) *Group 2: DL-segmentation:* The DL segmentation was performed for group 1 CT-data (n=757 individual muscle segmentation).

3) *Group 3: Randomly selected:* Extracted from the 5-fold ensemble used to create the full 3D muscle segmentation, we randomly selected axial CT slices at non-mid-vertebral levels from thoracic [T1: n=3, T2: n=3, T4: n=3, T5: n=7, T6: n=7, and T7: n=14] and lumber  [L1: n=6, L2: n=2, L3: n=8, L4: n=3, and L5: n=11], yielding 704 individual muscle segmentations.

4) *Group 4- External data: National Lung Screening Trial (NLST) dataset [4].* To demonstrate the model generalizability, we selected 30 subjects from the National Lung Screening Trial (NLST) [4], performed 3D DL muscle segmentation and evaluated the model performance for thoracic [T1: n=4, T2: n=3, T4: n=4, T5: n=4, T6: n=4, and T7: n=11] and 30  lumbar [L1: n=2, and L2: n=28] yielding 673 individual muscle segmentations. Table S.2 details the CT imaaging parameters for the NLST subjects [4].

**Table S.2** CT characteristics of the patients included from the Imaging Data Commons.

| CT Protocol parameters | Mean (SD) | Range |
| --- | --- | --- |
| Tube voltage (kVp) | 120.4 ( 2.9) | 120.4 ( 2.9) |
| Tube current (mA) | 107.5 (45.1) | 50 - 250 |
| X-Ray Exposure | 526.0 (523.4) | 25 -  1440 |
| In-Plane Pixel Size | 0.66 (0.64) | 0.55 - 0.78 |
| Slice Thickness (mm) | 2.21 (0.61) | 2.21 (0.61) |

References Cited.

1.	Allaire, B.T., et al., *Dependence of trunk muscle size and position on age, height, and weight in a multi-ethnic cohort of middle-aged and older men and women.* J Biomech, 2023. **157**: p. 111710.

2.	Robb, R.A., *The biomedical imaging resource at Mayo Clinic.* IEEE Trans Med Imaging, 2001. **20**(9): p. 854-67.

3.	Lee, J.-S., *Digital image smoothing and the sigma filter.* Computer vision, graphics, and image processing, 1983. **24**(2): p. 255-269.

4.	(ACRIN), L.S.S.g.L.a.t.A.C.o.R.I.N., *National Lung Screening Trial (NLST)* N.C.I. (NCI), Editor. 2009.
